# Estimating the size of COVID-19 epidemic outbreak

**DOI:** 10.1101/2020.03.28.20044339

**Authors:** Chakrit Pongkitivanichkul, Daris Samart, Takol Tangphati, Phanit Koomhin, Pimchanok Pimton, Punsiri Dam-O, Apirak Payaka, Phongpichit Channuie

**Affiliations:** Department of Physics, Faculty of Science, Khon Kaen University, Khon Kaen, 40002, Thailand; Department of Physics, Faculty of Science, Chulalongkorn University, Bangkok 10330, Thailand; School of Medicine, Walailak University, Nakhon Si Thammarat, 80160; Research Group in Applied, Computational and Theoretical Science (ACTS), Walailak University, Nakhon Si Thammarat, 80160, Thailand; School of Science, Walailak University, Nakhon Si Thammarat, 80160, Thailand; Plasmas and Electromagnetic Wave Science (PEwave) Center of Excellence, Walailak University, Nakhon Si Thammarat, 80160, Thailand; College of Graduate Studies, Walailak University, Nakhon Si Thammarat, 80160, Thailand

## Abstract

In this work, we analyze the epidemic data of cumulative infected cases collected from many countries as reported by WHO starting from January 21^st^ 2020 and up till March 21^st^ 2020. Our inspection is motivated by the renormalization group (RG) framework. Here we propose the RG-inspired logistic function of the form 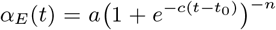 as an epidemic strength function with *n* being asymmetry in the modified logistic function. We perform the non-linear least-squares analysis with data from various countries. The uncertainty for model parameters is computed using the squared root of the corresponding diagonal components of the covariance matrix. We carefully divide countries under consideration into 2 categories based on the estimation of the inflection point: the maturing phase and the growth-dominated phase. We observe that long-term estimations of cumulative infected cases of countries in the maturing phase for both *n* = 1 and *n* ≠ 1 are close to each other. We find from the value of root mean squared error (RMSE) that the RG-inspired logistic model with *n* ≠ 1 is slightly preferable in this category. We also argue that *n* determines the characteristic of the epidemic at an early stage. However, in the second category, the estimated asymptotic number of cumulative infected cases contain rather large uncertainty. Therefore, in the growth-dominated phase, we focus on using *n* = 1 for countries in this phase. Some of them are in an early stage of an epidemic with an insufficient amount of data leading to a large uncertainty on parameter fits. In terms of the accuracy of the size estimation, the results do strongly depend on limitations on data collection and the epidemic phase for each country.

## INTRODUCTION & MOTIVATION

The coronavirus disease 2019 (COVID-19) caused by a novel coronavirus SARS-CoV-2 that emerged in the city of Wuhan, China, last year and has since expanded to a large scale COVID-19 epidemic and spread to all regions as reported by the World Health Organization (WHO) [1] causes a serious situation worldwide. Since the first reports of COVID-19, there are various attempts to estimate the final size of the spread mathematically. Among several approaches [2–9], the logistic growth models are very popular and useful to predict the final size of the epidemic [10, 11]. It is worth noting that the logistic function or logistic curve is a common “S” shape (Sigmoid curve). Logistic functions are often used in statistics and machine learning, medicine, chemistry, physics, material science, linguistics, and even agriculture. However its root somehow seems to be coincidentally originated from the high energy physics and condense matter physics’ point of view. In terms of the renormalisation group (RG) framework, we define the logistic growth function as an epidemic strength function allowing us to describe the spread of disease. Its derivative with respect to time yields a new quantity which can be interpreted as the beta function of an underlying microscopic model. This frame work was first proposed in Ref.[12] to describe the underlying dynamics of disease spread by invoking the usual logistic-growth model.

According to the reported data, a number of the COVID-19 infected cases grow up slowly at a very early stage of the epidemic spread. The next stage of growth is approximately exponential, then as saturation begins, the growth slows down to linear, and at maturity, the growth stops. Similar behaviour happens in various phenomena in physics, for example, the running coupling constants of particular theories describing phase transitions in high energy physics and in condensed matter physics. The renormalization group (RG) method is a successful framework, and is widely used to describe the dynamics of mentioned phenomena [13, 14]. We will use, in this work, the generalized characteristic function of the epidermic strength of the form

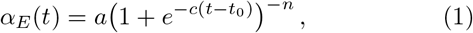

where *n* is an arbitrary constant, *a* is the curve’s maximum value, *c* is the logistic growth rate or steepness of the curve, and *t*_0_ can be thought of as a starting time at which *α*_*E*_(*t*_0_) = *a/*2^*n*^. It is worth noting that when *n* = 1 the strength function given in Eq.(1) is just a usual logistic one used in, for example, Refs.[10–12]. Here in the present work, we generalize such a function by considering its power-law form given in Eq.(1) and estimate the size of COVID-19 epidemic outbreak in terms of the state-of-the-art RG framework.

## RENORMALIZATION GROUP RECIPE

In this work, the epidemic strength function *α*_*E*_ in Eq.(1) will be identified as the number of the infected cases. This function is also known as the generalized logistic function. Moreover, the function *α*_*E*_ has many interesting features and it can be reduced to several useful function in physics such as Fermi-Dirac distributions [15] or running coupling constant of the four-fermion contact interaction [16] for *n* = 1 and the generalized Woods-Saxson nuclear potential [17] for *n* = *-*2 as well as the Starobinsky inflation potential [18] for *n* = − 2. Turning to the crucial part of this section, we will derive the beta function of epidemic strength coupling with the RG technique in the following and demonstrate how COVID-19 spread underlying the RG flow description. It is well known that the evolution of the *α*_*E*_(*t*) function is governed by the beta function. In the present work, we use *t* as a time of the evolution of the system instead of energy in high energy physics. The time scale in our work is related to the typical energy scale as *t* = ln *µ*_0_*/µ* in the standard RG approach where *µ*_0_ and *µ* are the energy scale limit of our interested and the running energy scale, respectively. The standard beta function of *α*_*E*_ is defined by

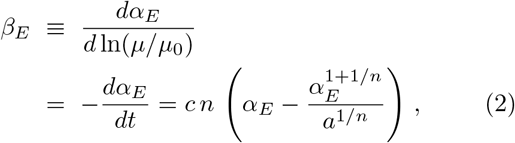

where *a, c* and *n* are arbitrary constants. An evolution (or RG flow) of the epidemic strength is controlled by the beta function in Eq.(2). The fixed points of the beta function are obtained by solving *β*_*E*_ = 0 which yield two fixed points in the RG equation as

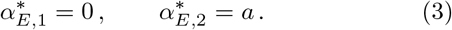

In order to explore the critical phenomena in the RG analysis, one needs to identify the stability and classify asymptotic properties of the fixed points from the the beta function. To do these, the slope of the beta function is very useful and convenient to study the stability for each fixed points. It reads,

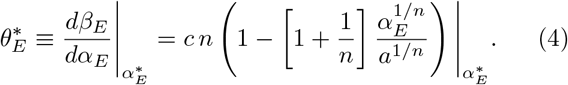

It is worth noting that the relative signs of the scaling exponent tell us about the fixed point stability. The plus sign reflects an unstable node while the negative value of the 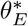 gives an attractor fixed point. Having use Eq.(4) for 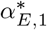,1 and 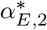, we obtain,

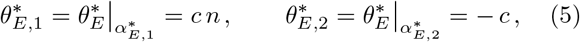

where the parameters *c* and *n* are positive constants and well defined for fitting procedure in the latter. On one hand, according to results in Eq.(5), the fixed point 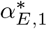 is the unstable node where this solution corresponds to the source or the origin of the dynamical system. On the other hand, the 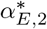 is the attractor fixed point which means the solution of the system asymptotically converging to this point. One may interpret the 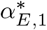 as the zero infected cases or starting point of the disease spreading while the maximum number of the infected cases and staying at this point under infinite time translation are represented by the 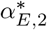 solution. As the results, we see that the spread of the COVID-19 can be represented by the running epidemic strength effect in the RG flow framework. Interestingly, moreover, the microscopic theoretical method in physics is suitable and useful to study the macroscopic phenomena such as the COVID-19 spread in the present study. Then, we would call the epidemic strenght function in Eq.(1) in this work as the RG-inspired model.

## FITTING RESULTS & INTERPRETATIONS

In the present analysis, we have collected data of the number of cumulative infections reported by WHO starting from January 21^st^ 2020 and up till March 21^st^ 2020, see [1]. The data are counted using the first day of an epidemic as day one such that the comparison between countries are feasible. We perform the non-linear least squares to fit with the 4-parameters model (*a, t*_0_, *c, n*) from Eq.(1). We are also interested in the case *n* = 1 where our results becomes the general model of population growth (S-curve). The uncertainty for model parameters is estimated using the squared root of the corresponding diagonal components of the covariance matrix. We carefully divide countries under consideration into 2 categories based on the estimation of the inflection point, *t*_0_, which is the point where the growth rate begins to decline, see FIG.1. We label any country where the current situation has already passed an estimated inflection point of the S-curve as currently in the *maturing phase*. A country in this phase will provide a good fit to all model parameters in our model. Whereas any country where the current situation has not reached an estimated inflection point yet is labelled as currently in the *growth-dominated phase*. Generally, a model fit of a country in this phase yields no reliable result. Only exponential function seems to fit a country in this phase.

**FIG. 1:**
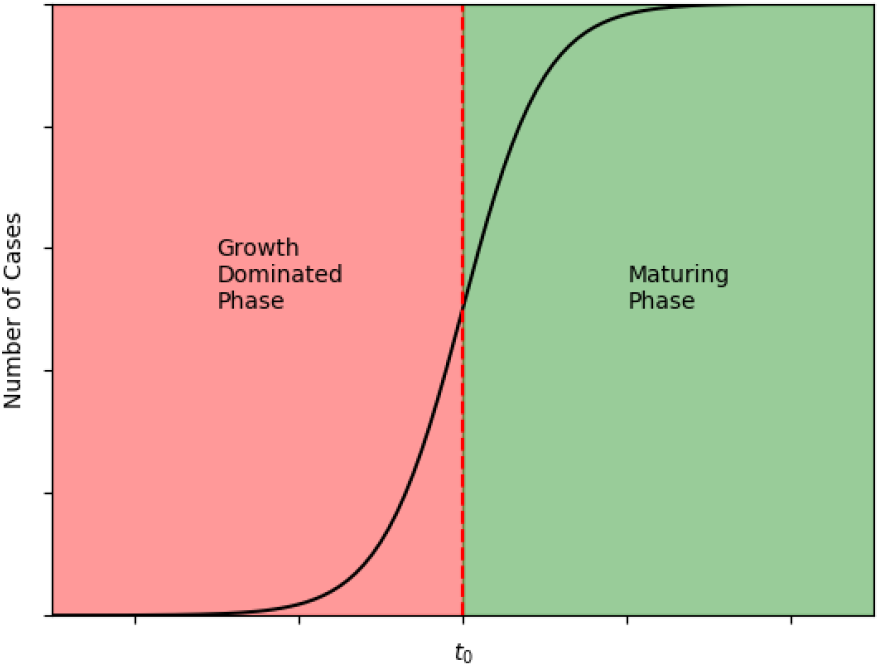
The classification of countries based on the inflection point. *t*_0_ is the midpoint of the logistic function (*α*_*E*_ (*t*_0_) = *a/*2) and the turning point where the growth rate begins to slow down.

### Maturing Phase

A clear example of countries beyond the inflection point is *China* where the outbreak was first reported. Another country where the outbreak has passed beyond the inflection point and the situation is now under control is *Korea*. The results of parameter fit for China and Korea are shown in Fig.2. In the China and Korea cases, The reported data show that the spread have reached the end point of the epidemic. This is the reason in our RG-inspired logistic growth model when both *n* = 1 and *n* ≠ 1 give very similar model parameters and very close prediction to each other. The value of root mean squared error (RMSE) from both models suggests that the RG-inspired logistic model with *n* ≠ 1 is slightly preferable.

**FIG. 2:**
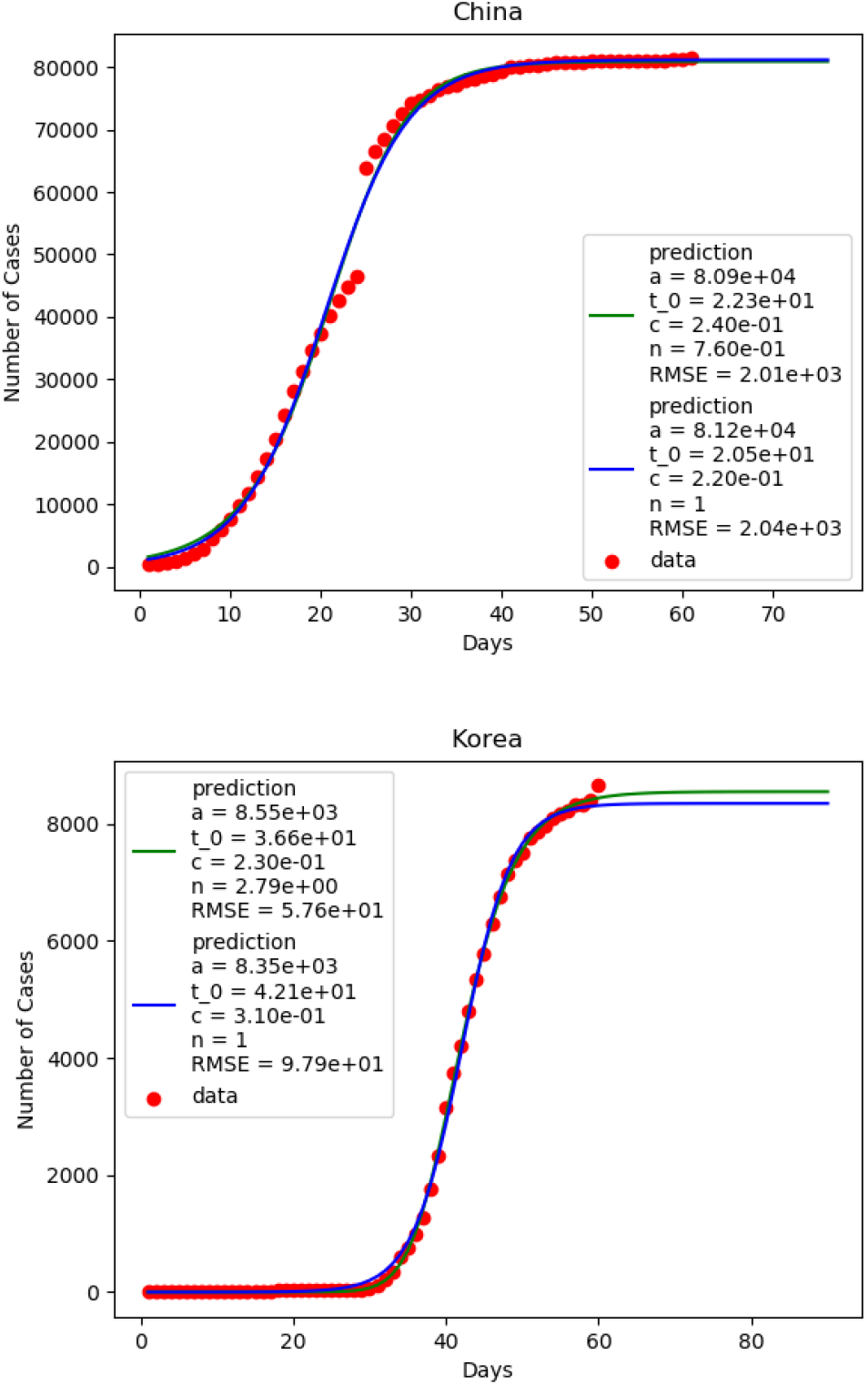
The parameter fit on data from China and Korea. The green line and blue line are the best fit from RG-inspired logistic model and logistic model respectively. Both models are very close in predicting the size of the epidemic.

Another set of countries where the situation has already passed the inflection point but has not reached the plateau yet are *Austria, Belgium, Japan*, and *Norway*. The results of fitting are shown in Fig.3 for Austria and Japan.

**FIG. 3:**
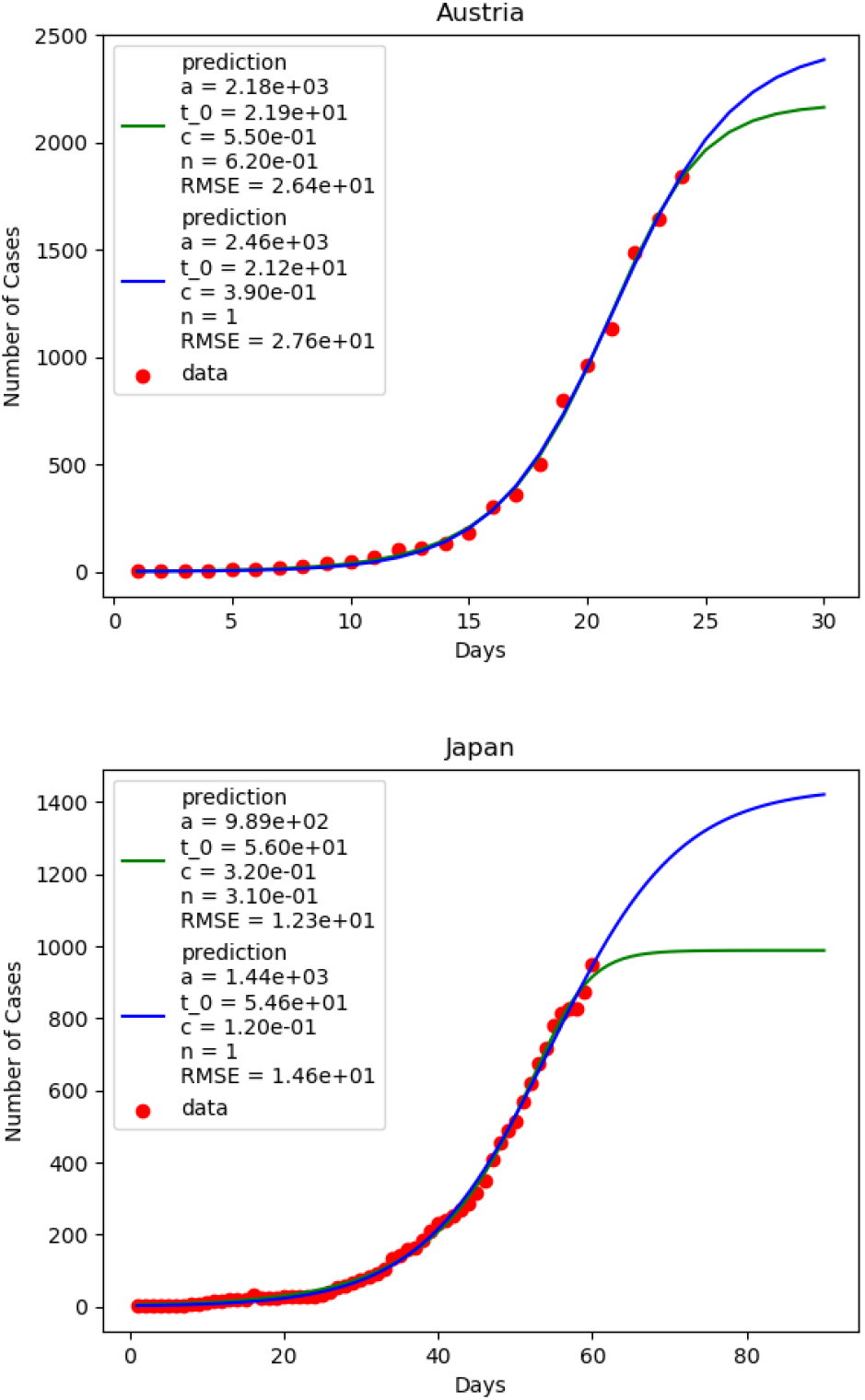
The parameter fit on data from Austria and Japan. The green line and blue line are the best fit from RG-inspired logistic model and logistic model respectively.

Compared with *n* = 1, the RG-inspired logistic model with *n* ≠1 is again a slightly better fit to data, and the prediction of the size of the outbreak is lower than that of *n* = 1. The values of the model parameters with estimated errors are given in TABLE I. The values of *n* in RG-inspired logistic model represent asymmetry in the logistic function. When *n* < 1, initial phase of the epidemic is slower than that observed in the normal logistic function (*n* = 1) see Fig.4.

**TABLE I:** The table shows the best-fitted values of parameters (*a, t*_0_, *c, n*) for each country categorized in the maturing phase. Here we observed that *n* < 1 in all cases implying that initial phase of the epidemic is slower than that of the *n* = 1 case, a normal logistic function.

| Country | $a$ | $t_0$ (days) | $c$ (day$^{-1}$) | $n$ |
| --- | --- | --- | --- | --- |
| China | 80914±448 | 22.3±1.1 | 0.24 ± 0.02 | 0.76 ± 0.13 |
| Korea | 8548±35 | 36.6±0.9 | 0.23 ± 0.01 | 2.79 ± 0.43 |
| Austria | 2179±192 | 21.9±0.4 | 0.55 ± 0.15 | 0.62 ± 0.22 |
| Belgium | 2591±806 | 43.5±1.0 | 0.36 ± 0.20 | 0.83 ± 0.61 |
| Japan | 988±37 | 56.0±0.3 | 0.32 ± 0.05 | 0.31 ± 0.05 |
| Norway | 1725±124 | 18.5±1.0 | 0.41 ± 0.10 | 0.84 ± 0.33 |

**FIG. 4:**
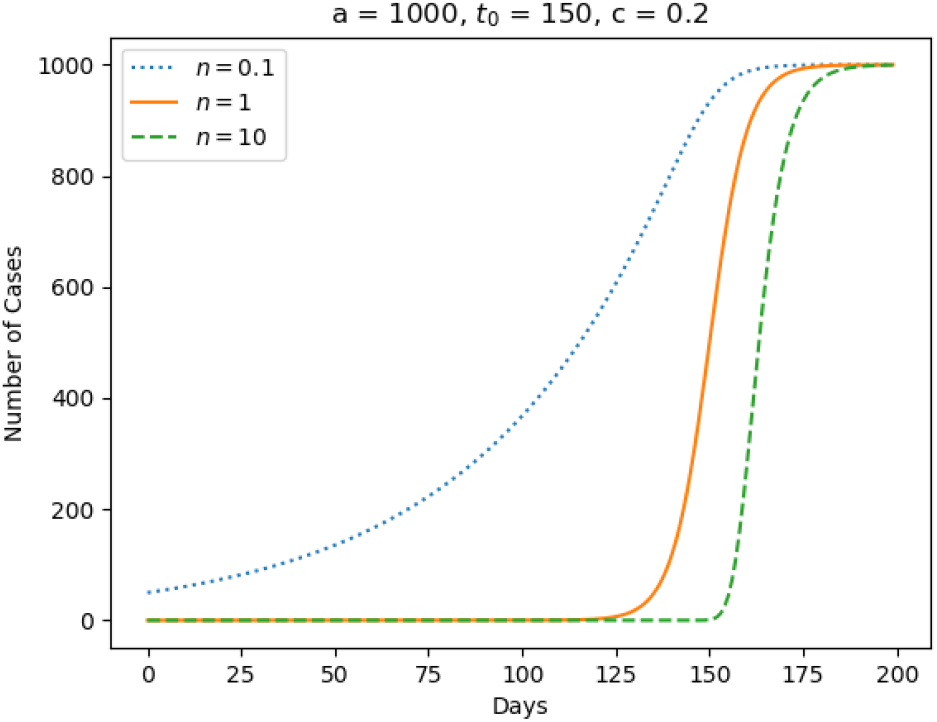
The plots demonstrate the effect of *n* on the population growth. *n >* 1 suggests a growth rate higher than typical logistic function (*n* = 1) whereas *n* < 1 suggests a slower growth.

China, Japan, Austria, Belgium and Norway are fitted into this group. These countries have evidently chosen a relatively strong measure in containing and mitigating the spread of the virus since an early stage. We argue that this value is a reliable indication of an early stage effort on controlling the epidemic. Korea, on the other hand, has the value of *n* = 2.79*±*0.43 suggesting that the spread of the virus in an early stage was much quicker than the normal logistic function (*n* = 1). This coincides with the super-spreader event [19] that happened in Korea on 20 February 2020. Using *n* as a characterisation on asymmetry, we in the next subsection proceed the estimation analysis on countries in growth-dominated phase.

### Growth-Dominated Phase

One of the countries which has not reached the expected inflection point yet is *Italy*. The results of fitting with data for various values of *n* is shown in Fig.5. Our results suggest that the early stage of epidemic in Italy is not in a controlled stage since the data in early days fit better with a higher value of *n*. However, we do not pursue the exact value of *n* of the country before the expected inflection point because of the large uncertainty coming with other parameters especially the size of the epidemic, *a* which we will discuss later.

**FIG. 5:**
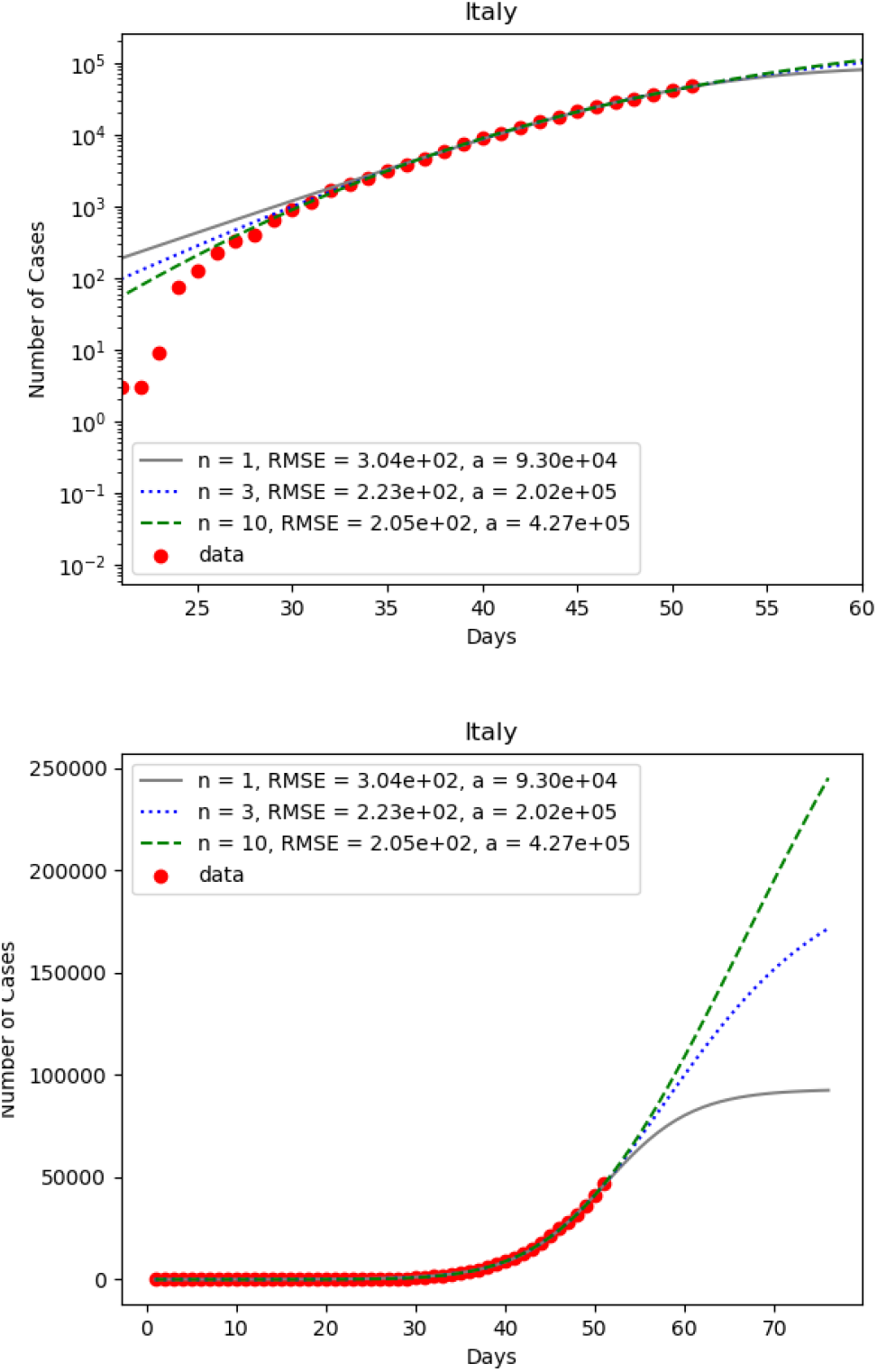
The parameter fit on data of cumulative infected cases of Italy for *n* = 1, 3, 10.

Instead, we first focus on using *n* = 1 for countries in this phase. Some of them are in an early stage of epidemic with insufficient amount of data leading to a large uncertainty on parameter fits. Many of them have an official number of infected cases that might be under investigated due to limited resources in testing kits. We will focus on 5 European countries (*Italy, Spain, France, Germany, and Switzerland*) since their outbreaks have been happening for a while and their health system are somehow capable of reporting data. Examples of the results from parameter fit with *n* = 1 are given in Fig.6 where the error of parameter fits are displayed in TABLE II.

**TABLE II:** The table shows the best-fitted values of parameters (*a, t*_0_, *c*) for each country categorized in the growth-dominated phase. Here the results are of the parameter fit with *n* = 1. In this case, we observed that the best fits still contain higher uncertainty compared with the countries in the maturing phase.

| Country | $a$ | $t_0$ (days) | $c$ (day<sup>-1</sup>) |
| --- | --- | --- | --- |
| Italy | $92968 \pm 4379$ | $51.1 \pm 0.4$ | $0.206 \pm 0.004$ |
| Spain | $35267 \pm 1723$ | $49.2 \pm 0.3$ | $0.325 \pm 0.008$ |
| France | $22894 \pm 1077$ | $56.5 \pm 0.3$ | $0.262 \pm 0.004$ |
| Germany | $52579 \pm 17033$ | $58.6 \pm 1.9$ | $0.242 \pm 0.010$ |
| Switzerland | $6692 \pm 1209$ | $23.2 \pm 1.2$ | $0.301 \pm 0.026$ |

**FIG. 6:**
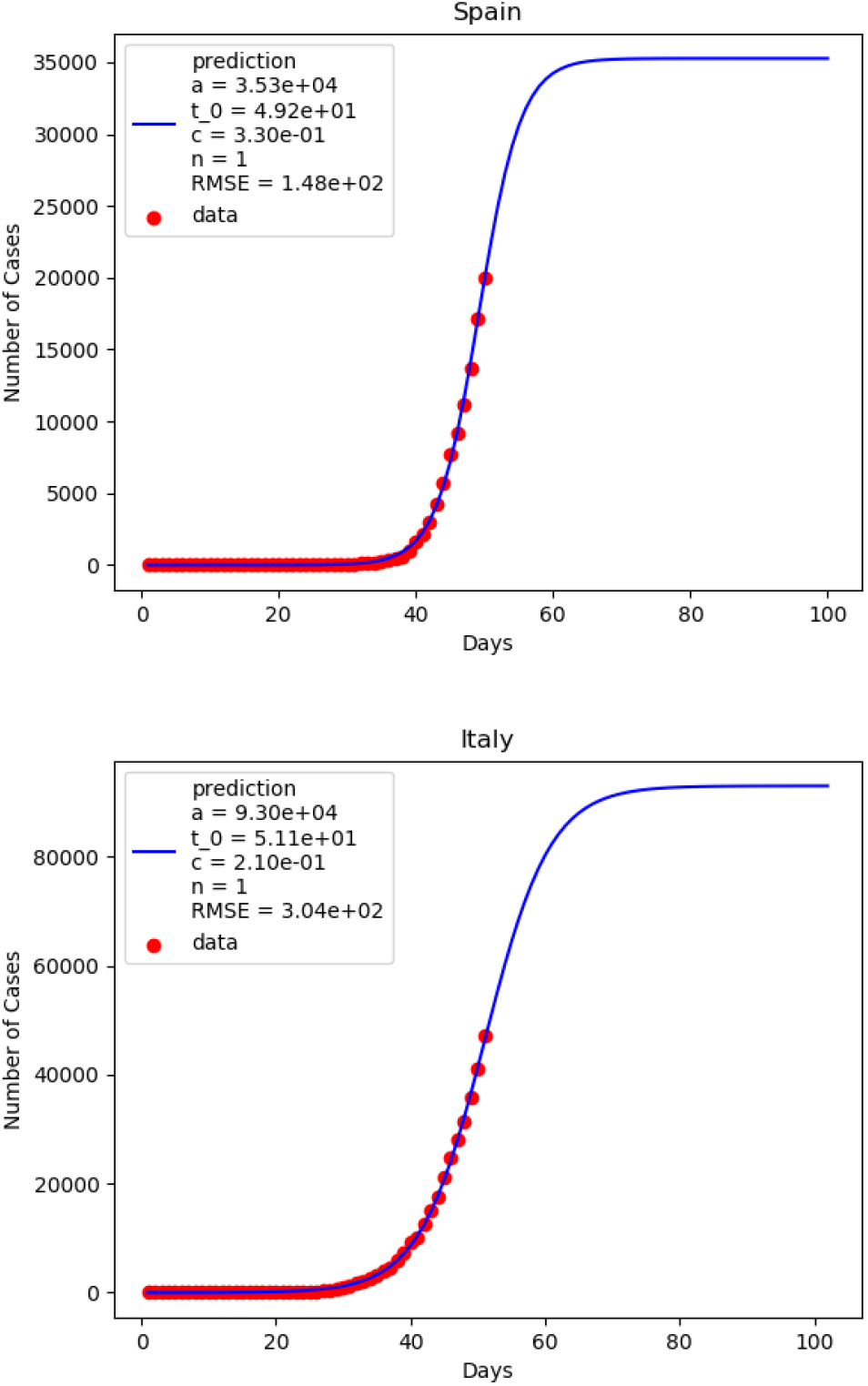
The parameter fit on data of cumulative infected cases of Spain (upper panel) and Italy (lower panel) for *n* = 1.

To complete the discussion on the estimation of epidemic size for countries in growth-dominated phase, we find the best fit for each value of *n* ranging from 1-5 and provide the multiplying factor *a/a*_0_ where *a*_0_ is the prediction of the size of each epidemic from normal logistic function with *n* = 1 (see the second column in TABLE II). The results are given in FIG.7. The multiplying factor can be interpreted as an uncertainty in the final size of each epidemic. In order to provide a reliable prediction of model, we need more data from these countries.

**FIG. 7:**
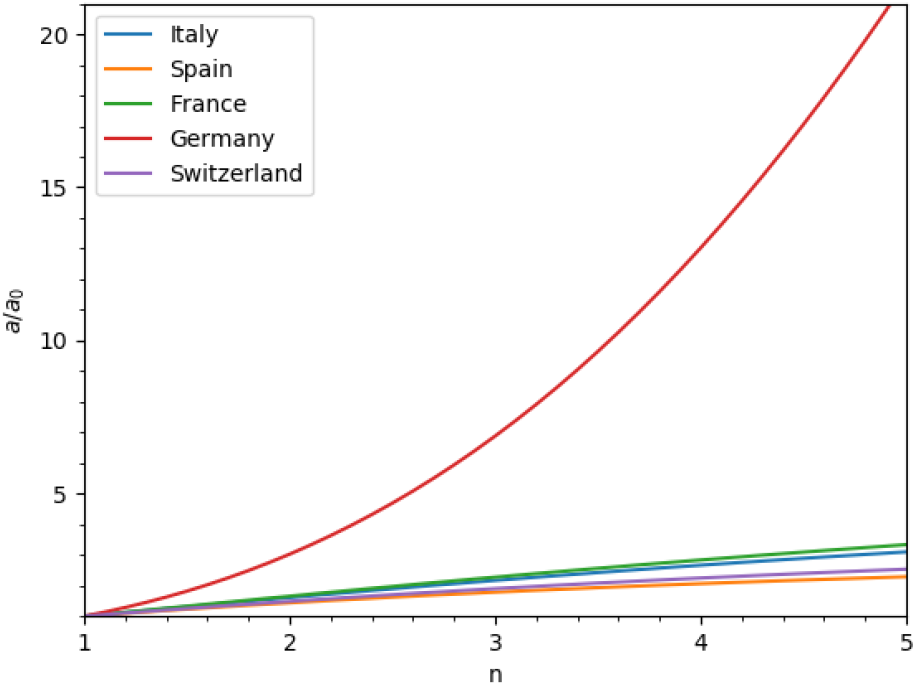
The multiplying factor *a/a*_0_ as a function of *n* of 5 European countries. We observed that up till *n ≤* 5 the factor *a/a*_0_ is not greater than 2 apart from Germany. This factor is useful for the estimation of the size of epidemic.

## DISCUSSION & CONCLUSION

In this section, we concluded our findings on analyzing the epidemic data of cumulative infected cases from many countries as reported by WHO. In terms of a renormalisation group approach, we considered the RG-inspired logistic growth function, a.k.a. the power-law logistic growth function. The non-linear least squares regression is performed to obtain parameters from the model. We computed the uncertainty for model parameters using the squared root of the corresponding diagonal components of the covariance matrix. We carefully divided countries under consideration into 2 categories based on the estimation of the inflection point, *t*_0_: the maturing phase and the growth-dominated phase. We found that the outbreak has happened in a large scale and passed beyond the inflection point include China and Korea, while countries where the situation has already passed the inflection point but has not reached the plateau yet are Austria, Belgium, Japan, and Norway with Japan being close to the asymptotic number of infected cases. We observed that long-term estimations of all countries in the maturing phase for both *n* = 1 and *n* ≠1 are close to each other. Based on the values of root-mean-square error, the RG-inspired logistic model is slightly preferable. Our further investigation shows that *n* characterizes an early stage of the epidemic. *n* < 1 represents countries with a strong measure at the beginning of the outbreak, where *n >* 1 indicates an uncontrollable event.

In the second category, we consider all countries that the cumulative number of infected cases does not reach the expected inflection point, yet. The European countries that we consider fall in this category include Italy, Spain, France, Germany, and Switzerland. Due to the large uncertainty coming from other parameters especially the size of the epidemic, *a*, in this category we focus on using *n* = 1 for countries in this phase. We estimate the asymptotic number of cumulative infected cases of countries in the growth-dominated phase. We also provide an estimate of the epidemic size as a function of *n*. However, in order to obtain a reliable prediction of model, we need more data from these countries in the second category. In conclusion, we provide an alternative way to describe the spread of COVID-19 using the renormalization group method originated from physics’ point of view.

## Data Availability

The authors have declared no competing interest.

